# Evaluation of SSI risk prediction model after spinal surgery: A systematic review and critical appraisal

**DOI:** 10.64898/2025.12.15.25342321

**Authors:** Mohan Du, Ru Zhou, Hepeng Du, Xiu Li, Liping Ying

**Author notes:** Corresponding Author: Liping Ying.

## Abstract

This study aimed to systematically review and critically evaluate the risk of bias and applicability of surgical site infection (SSI) risk prediction models after spinal surgery. China National Knowledge Infrastructure, Wanfang Database, China Science and Technology Journal Database (VIP), SinoMed, PubMed, Web of Science, The Cochrane Library, Cumulative Index to Nursing and Allied Health Literature, and Embase were searched from inception to April 10, 2025. The prediction model risk of bias assessment tool-artificial intelligence (AI) and transparent reporting of a multivariable prediction model for individual prognosis or diagnosis-AI were used to assess the quality of the included studies, and RevMan software was used to perform a meta-analysis of the odds ratio values for certain model predictors. A total of 37 studies were included, identifying 43 predictive models. The incidence of SSI after spinal surgery ranged from 1.5% to 50%. Among these, 11 studies focused solely on model development, 4 studies included external validation, 22 studies were only internally validated, and 1 study was both internally and externally validated. The area under the curve values ranged from 0.610 to 0.991. The meta-analysis of high-frequency predictors identified statistically significant factors, including diabetes, age, surgery duration, albumin, body mass index, drainage time, smoking history, and American Society of Anesthesiologists score. All studies were rated as having a high risk of bias, primarily due to poor reporting related to study participants and the analysis domain. The evaluation using the prediction model risk of bias assessment tool indicated a considerable risk of bias in current predictive models for postoperative SSI after spinal surgery. Although the predictive model for SSI after spinal surgery is generally acceptable, most studies have methodological flaws. Moreover, studies with larger sample sizes and multicenter external validation are necessary to enhance the robustness of predictive models.

## 1 Introduction

Orthopedics is the fourth most common type of surgery worldwide. Surgical site infection (SSI) is the most common healthcare-associated infection globally, leading to a high incidence of severe complications (*Centers for Disease Control and Prevention*, 2024) and posing a significant burden on healthcare systems (Patel et al., 2017). The SSI is among the most serious complications after spinal surgery (Lee et al., 2014). It can result in adverse surgical outcomes, such as fixation failure, osteomyelitis, prolonged hospital stay, and increased mortality (Janssen et al., 2019; Pesenti et al., 2018), along with high medical costs, ranging from $15,800 to $43,900 (Manian, 2014; Whitmore et al., 2012; Yeramaneni et al., 2016). Perioperative measures are essential to reducing the risk of SSI. SSI prophylaxis consists of three phases: preoperative, intraoperative, and postoperative. Preoperative and intraoperative measures used in spinal surgery include antimicrobial prophylaxis, preoperative disinfection baths, and intraoperative skin disinfection preparations. However, the postoperative phase is often neglected due to preconceived notions about surgery. Using antiseptics and antiseptic drugs in spinal surgery is often overlooked, and evidence supporting their effectiveness in SSI prevention remains limited (Manian, 2014). Consequently, assessing the risk of SSI and prescribing preventive measures accordingly are extremely important to prevent SSI and avoid potentially devastating consequences.

SSI after spinal surgery affects patient prognosis, and effective management of SSI is crucial. Early identification of risk factors is essential for implementing tailored prevention strategies for specific populations. The risk prediction model plays a vital role in this process. These models utilize multiple predictors—often in the form of regression equations, nomograms, or artificial intelligence (AI)-based algorithms—to estimate the probability of SSI occurrence. Although a systematic review of the SSI prediction model after spinal surgery was conducted (Lauinger et al., 2024), it lacks updates based on recent studies and does not reflect the latest methodological guidance and quantitative evaluation. A comprehensive evaluation is essential for selecting the most suitable prediction models for medical staff.

This study aimed to identify published studies on the development or validation of predictive models for SSI after spinal surgery and to critically evaluate existing models, informing clinical practice and guiding future research. Additionally, this study evaluates the use of a prediction model risk of bias assessment tool (PROBAST)-AI and transparent reporting of a multivariable prediction model for individual prognosis or diagnosis (TRIPOD)-AI guidelines in assessing the quality of machine learning (ML)-based predictive models, reflecting the growing role of AI technologies in predictive modeling. Consequently, this review aims to systematically evaluate the performance and characteristics of prediction models for SSI after spinal surgery, identify the best-performing models, and critically evaluate their risk of bias and clinical applicability. It also highlights opportunities for future research and aims to strengthen the clinical application, providing a reference for future model development. **2 | Materials and Methods**

This systematic review was conducted following the guidelines outlined in the Preferred Reporting Items for Systematic Reviews and Meta-analysis (PRISMA) reporting guideline (Page et al., 2021). The study protocol was registered in the Prospero International Prospective Register of Systematic Reviews (CRD42024516 854).

### 2.1 Search Strategy

To ensure a comprehensive search, we targeted databases in both English and Chinese languages, acknowledging the broad population coverage and language diversity in these regions. The databases we searched included PubMed, Web of Science, EBSCO CINAHL Plus, Embase, Cochrane Library, China National Knowledge Infrastructure, Wanfang, Chinese Biomedical Literature Database, and Weipu. The search spanned from the inception of the databases until April 10, 2025. We employed a robust search strategy combining Medical Subject Headings and free-text terms within the titles, abstracts, and keywords. Furthermore, we utilized a retrieval filter focusing on prediction models, complemented by manual and citation retrieval methods. Our basic search incorporated keywords such as “spine surgery,” “spinal fusion,” “spine fractures,” “surgical wound infections,” “SSI,” “rule,” “predictor,” “model,” “risk assessment,” “risk score,” and “algorithm.” For a detailed overview of our search strategies, refer to Supplementary Appendix 1.

The review was structured using the PICOTS framework, as advised by the Critical Appraisal and Data Extraction for Systematic Reviews of Prediction Modeling Studies (CHARMS) checklist (Moons et al., 2014), outlining the essential elements of our study as follows:

**P** (Population): Adults aged 18 years and older.

**I** (Intervention model): Risk prediction models for SSI after spinal surgery that included predictors ≥ 2.

**C** (Comparator): No comparator model was required.

**O** (Outcome): The primary outcome was the occurrence of SSI (not its subgroups).

**T** (Timing): Predictions based on preoperative, intraoperative, or postoperative factors.

**S** (Setting): Clinical settings where risk prediction models are used to personalize SSI risk estimates and support preventive strategies.

### 2.2 Inclusion and Exclusion Criteria

The inclusion criteria were as follows: (1) Studies involving adults diagnosed with post-spinal surgery SSI; (2) studies addressing prediction models for SSI after spinal surgery, providing detailed descriptions of the models’ construction, validation, and evaluation processes; (3) study designs including cohort studies, cross-sectional studies, randomized controlled trials, and case-control studies; (4) studies published in either Chinese or English.

The exclusion criteria included the following: (1) Informally published literature, such as conference abstracts, news reports, and preprints; (2) studies with limited availability of methodological data; (3) research conducted at the cellular or molecular levels; (4) Prediction models based solely on virtual or simulated data; (5) models with fewer than two predictive variables in the final model; (6) outcomes limited to SSI subgroups; (7) inability to access the full text of the literature.

### 2.3 Literature Screening and Data Extraction

Two reviewers independently screened the search results. The eligibility of full-text reports was assessed, and any disagreements were resolved through discussion or with the involvement of a third reviewer. Data extraction was guided by the CHARMS checklist. The collected data included key characteristics of all included studies, such as the first author, year of publication, country of research, study design, study population, data source, and percentage of events. Details regarding the predicted outcomes were also captured, including outcome definitions and information about the development of prediction models. This encompassed the number of candidate variables, methods for processing continuous variables, sample size, number of outcome events, number of missing data entries, handling methods, details of model establishment, and variable selection methods. Besides, insights into model performance and predictive factors were documented, encompassing model performance evaluation, calibration methods, final predictive factors, and model presentation.

### 2.4 Assessment of Study Quality

Three researchers (RZ, XL) independently assessed potential sources of bias using the PROBAST-AI (Moons et al., 2025). PROBAST-AI assesses the risk of bias across four key domains: participants, predictors, outcome, and analysis. Applicability is evaluated across three domains: participants, predictors, and outcome. The assessment within each domain involves responding to signaling questions with options of “yes,” “probably yes,” “no,” “probably no,” or “no information.” The risk of bias and applicability are categorized as low, high, or unclear. A study’s overall risk of bias is considered low if all domains are rated as low, high if any domain is rated high, and unclear if one or more domains are rated as unclear while the remaining domains are rated low.

To assess reporting transparency, we applied the TRIPOD-AI checklists (Collins et al., 2024). This tool includes 25 checklist items for studies that involve both model development and validation, and 31 items for studies exclusively focused on either model development or validation. The TRIPOD-AI score was calculated as the percentage of checklist items fulfilled by each study.

### 2.5 Data Analysis

During the analysis phase, we systematically compared the modeling methodologies and selected the highest-performing model for reporting. Each unique combination of temporal predictors was treated as an independent model. For predictors appearing in over five studies, we conducted odds ratio (OR) meta-analyses using RevMan software.

Heterogeneity was assessed using the I^2^ metric and the Cochran Q test to determine whether a fixed-effect or random-effects model was appropriate. GraphPad Prism software (version 10.0) was used to handle the area under the curve (AUC) values. Microsoft Excel was used to generate visual representations of predictor distributions and to evaluate bias risk.

## 3 Results

### 3.1 Study Selection

Initially, 7,067 records were retrieved through the systematic search. After removing 1,158 duplicates, the titles and abstracts of the remaining 5,909 references were screened for eligibility. Moreover, four additional studies were identified through the citation retrieval of related systematic reviews, and full-text assessments were conducted on 52 studies to determine their compliance with the predefined inclusion criteria. Among these, six studies were excluded because they did not focus solely on risk factors. Similarly, three studies were excluded due to content inconsistencies with the review; one was excluded because it used the same samples and data, and four were excluded because they lacked sufficient information. Consequently, 37 studies were included in our systematic review. The PRISMA flowchart outlining the search strategy and selection process is provided in Figure 1(Flow chart).

**Figure 1:**
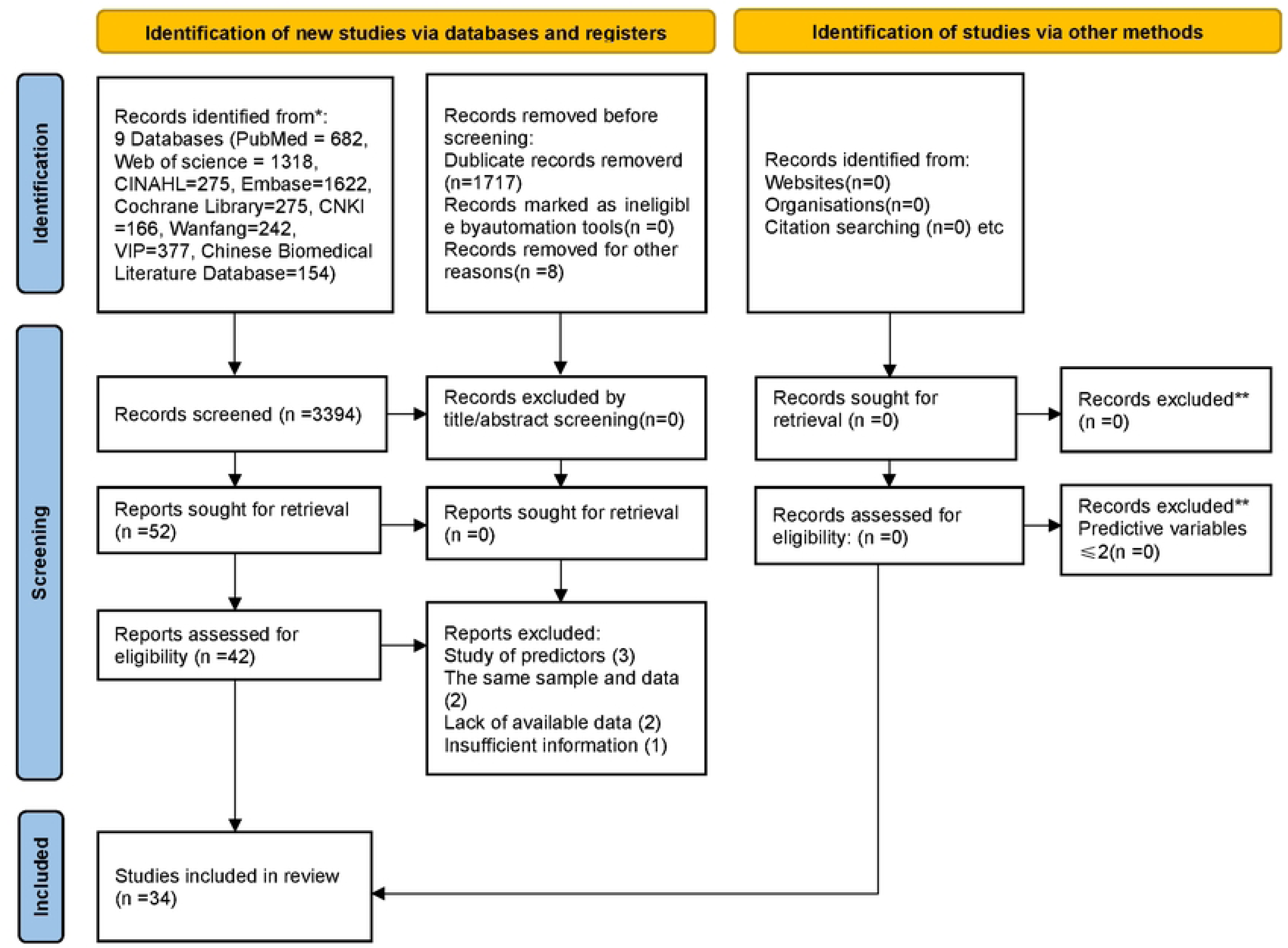
PRISMA 2020 flow diagram for updated systematic reviews which included searches of databases and registers only

### 3.2 Characteristics of the Included Studies

The design and participant characteristics of the 37 included studies are provided in Table 1(Basic characteristics of included studies). Approximately 90% of the studies were published within the last five years. The reported rates of SSI after spinal surgery varied widely, ranging from 1.5% to 50.0%. Sample sizes ranged from 98 to 4,046 participants. Geographically, 27 studies were conducted in China, including 7 published in Chinese, 2 in Russia, 5 in the USA, 2 in the Netherlands, and 2 in Japan. Among the included studies, 36 were retrospective, including 2 multicenter studies, and 2 were case-control studies. The Centers for Disease Control and Prevention criteria were the most commonly used diagnostic standard for SSI, followed by criteria from some tertiary hospitals in China.

Key features related to the construction of prediction models are detailed in Table 2(The basic characteristics of constructing forecasting model). Among the included studies, three focused solely on external model validation, while the remainder were dedicated to model development. The number of candidate predictors considered in the models ranged from 11 to 43, and the events per variable (EPV) ratio varied from 0.380 to 165.625. Most models employed logistic regression (LR) to identify independent predictors of SSI after spinal surgery. Furthermore, two models used a single ML technique, and five used multiple ML techniques. Random forest was the most frequently used ML technique, appearing in four studies, followed by decision tree and support vector machine, each used in three studies. Among the 37 papers analyzed, a total of 43prediction models were collectively constructed. Notably, 37 optimal models were identified across 38 studies, with two studies each presenting a single model. Common presentation methods included nomograms, scoring systems, and equation-based models.

### 3.3 High-frequency Predictors and Meta-analysis

The final model of the model development study comprises 2–8 factors, totaling 65 predictive elements. These factors include preoperative, intraoperative, and postoperative variables. The frequency distribution of the predictors included in the final model is depicted in Figure 3, with each predictor appearing at least twice. Among these predictors, diabetes, age, and time to surgery were the most commonly used, appearing in 16 and 13 models, respectively. Other commonly used predictors included albumin and the body mass index (BMI), each appearing in 11 models. Drain duration, blood loss, and the number of fused segments appeared in 9, 8, and 6 models, respectively, while smoking history and American Society of Anesthesiologists (ASA) classification were each included in 7 models. For details, please refer to Supplementary Appendix 2.

To further assess the significance of high-frequency predictors, a meta-analysis was conducted by extracting the OR values and confidence intervals (CIs) for each variable. The forest plot results indicated that there was no heterogeneity in the smoking history across studies (I^2^ < 50%, *P* > 0.1), allowing for the use of a fixed-effects model. Other factors indicated heterogeneity (I^2^ > 50%, *P* < 0.1), necessitating the use of a random-effects model. The statistically significant predictors of SSI after spinal surgery include diabetes (OR: 3.22, 95% CI: 2.01–5.16), age (OR: 1.05, 95% CI: 1.01–1.08), surgery duration (OR: 2.77, 95% CI: 1.57–4.90), albumin (OR: 0.86, 95% CI: 0.76–0.98), BMI (OR: 1.50, 95% CI: 1.17–1.92), drain duration (OR: 1.80, 95% CI: 1.32–2.45), smoking history (OR: 1.50, 95% CI: 1.17–1.92), and ASA (OR: 2.25, 95% CI: 1.79–2.82). In the meta-analysis, blood loss (OR: 1.00, 95% CI: 0.99–1.01) indicated statistical non-significance. The forest plot is depicted in Figure 2(meta-analysis) , and detailed OR values for each study are presented in Appendix 3—model development and validation performance.

**Figure 2:**
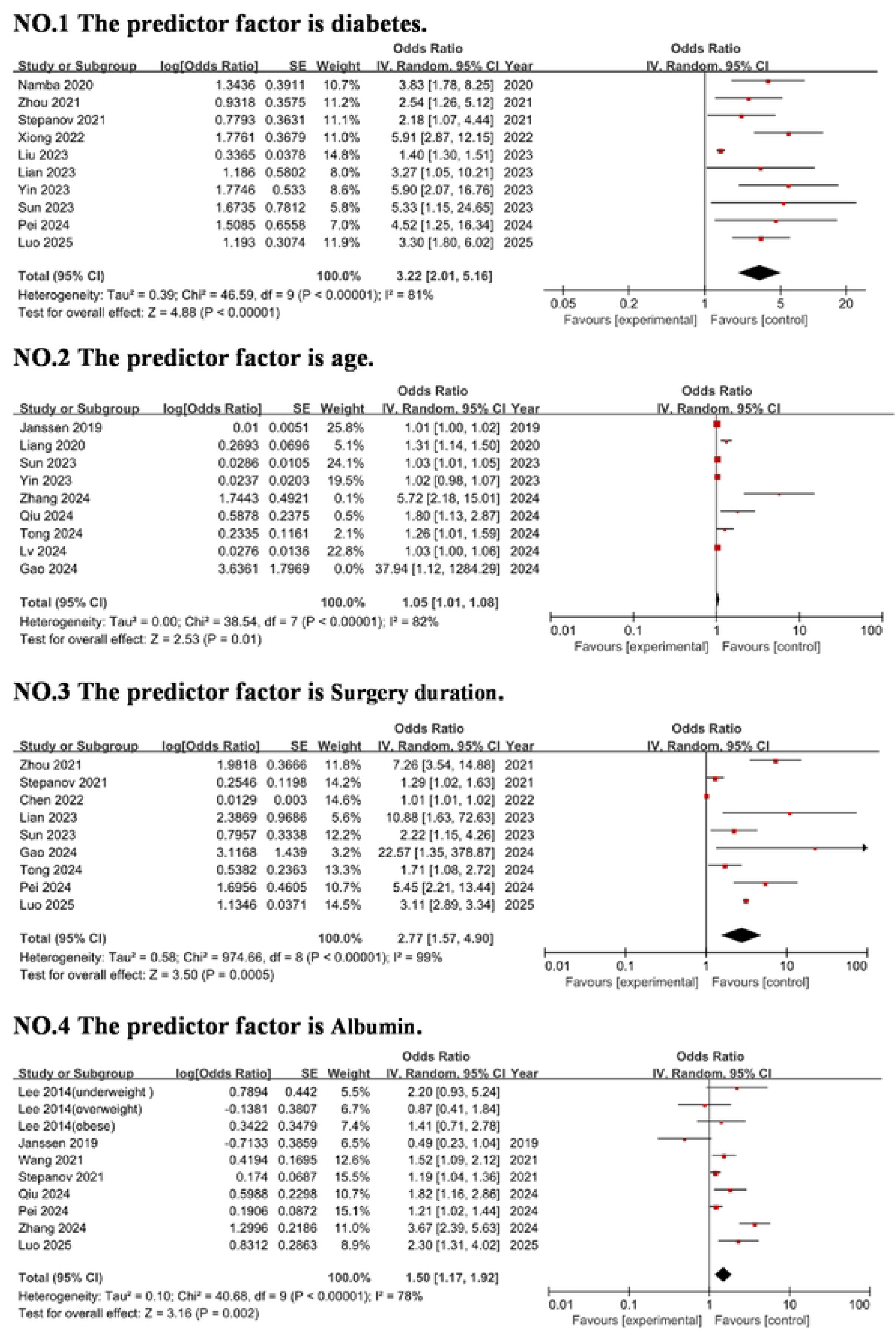
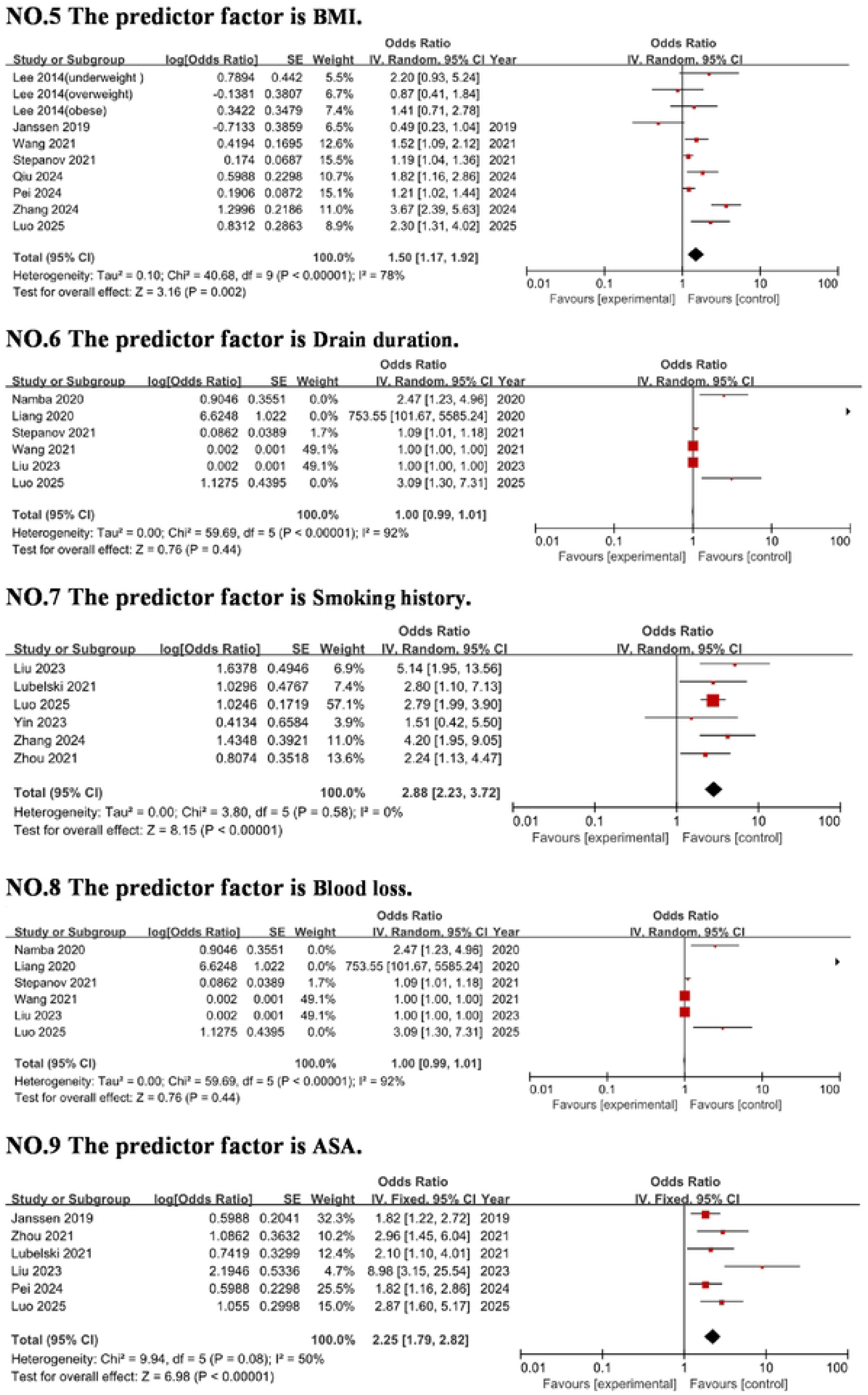
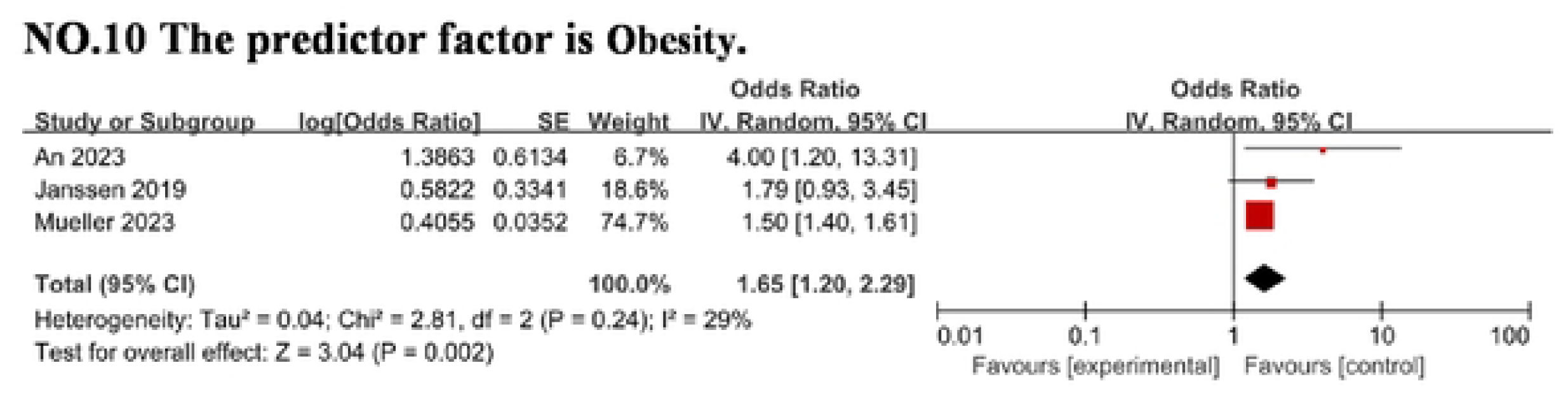
Meta-analysis of high-frequency predictors

**Figure 3.**
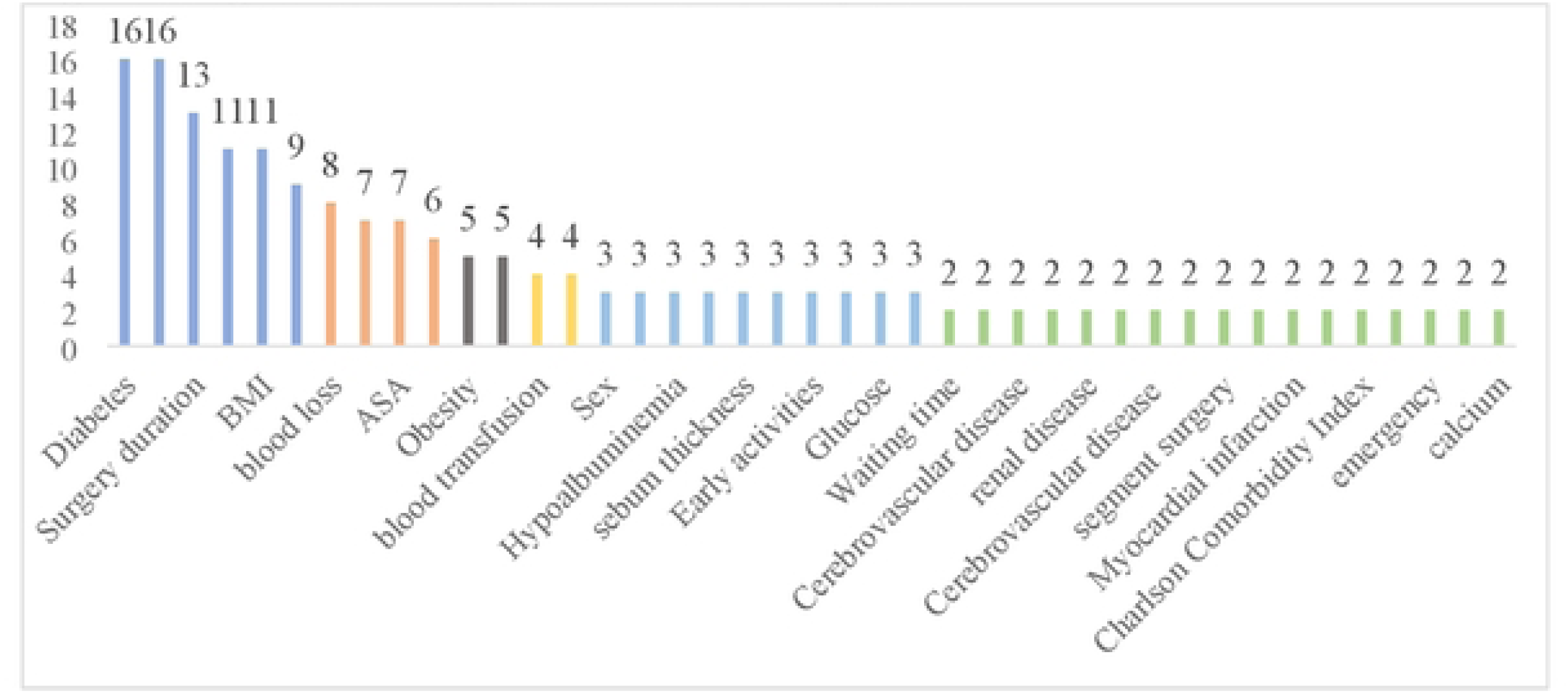
High frequency predictor frequency table

The reported AUC or c-statistic values ranged from 0.610 to 0.991, as detailed in Figure 4(Visualization of AUC value of prediction model). Calibration was evaluated in 24 models (64%), with the calibration curve being the primary method employed. Using the Hosmer–Lemeshow goodness-of-fit test for calibration assessment was observed in only eight studies. In the included studies, most models were validated both internally and externally. Four studies performed external validation, while 22 studies performed internal validation only. Notably, the model developed by Liang et al. was subjected to internal and external validation (Liang, 2020). The reported AUC or C statistic ranged from 0.891 to 0.991 in the internal validation set and from 0.610 to 0.991 in the external validation set. The visualization of the model AUC value is depicted in Figure 4.

**Figure 4.**
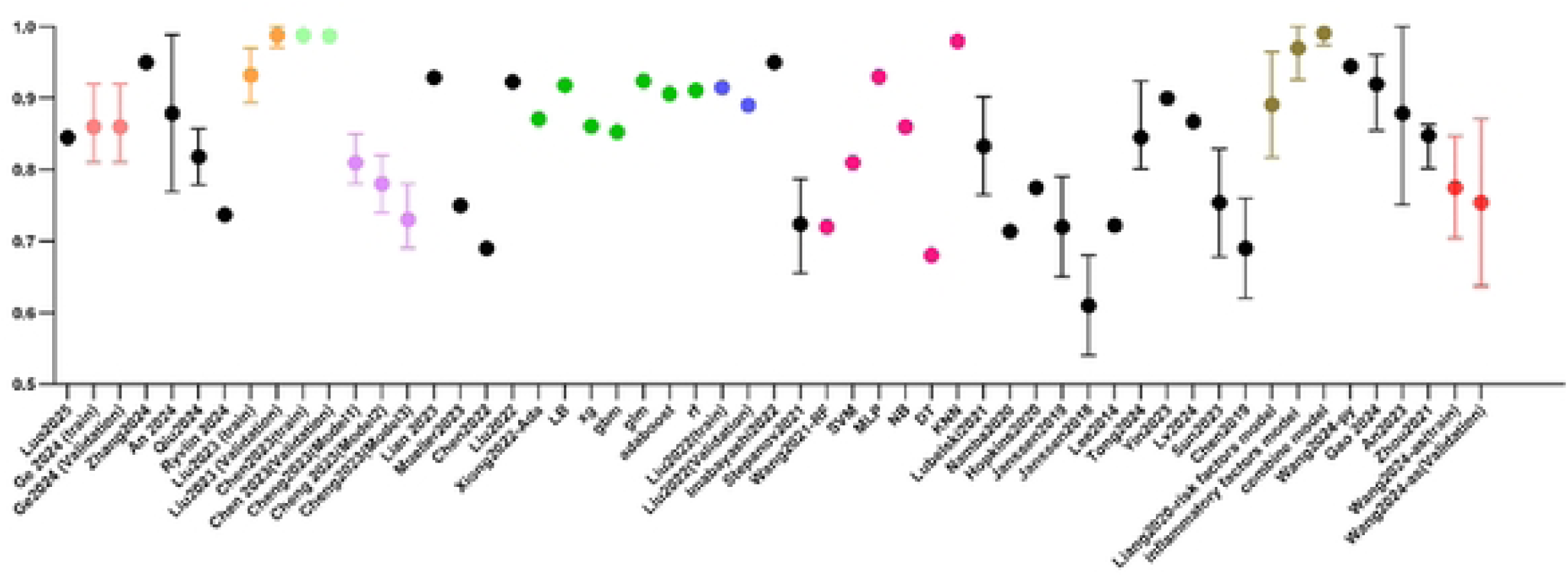
Visualization of AUC value of prediction model

### 3.4 Risk of Bias and Transparent Reporting

The methodological quality and reporting transparency of the 37 identified studies were summarized in Table 3(Quality Assessment). All studies were assessed as having a high risk of bias. Contributing factors included the retrospective nature of the study designs, lack of reported inclusion and exclusion criteria, absence or inappropriate reporting of predictor screening methods, insufficient reporting or improper handling of missing data, and the unreported assessment of whether sample sizes were adequate. In the subject domain, most studies were rated at high risk of bias due to their reliance on retrospective data, and 15 studies did not report inclusion and exclusion criteria. In the area of predictors, most studies do not explicitly assess time and are at high risk of bias, requiring specialized evaluation. Moreover, the lack of standardized definitions for predictors and the presence of missing blinded information in retrospective studies contributed to bias.

In the outcome domain, 11 studies had a high risk of bias. Issues include deviations from standard outcome definitions and overlapping predictors and outcomes. The 10 studies did not specify an appropriate time interval between the assessment of the predictor and the ascertainment of the outcome. In the area of analysis, 28 studies did not explain how to determine sample size, and nearly one-third of the studies transformed continuous variables into categorical variables, which may reduce the model’s accuracy. The treatment of missing data was not clarified in 28 studies, increasing the risk of bias. The treatment of missing data varied, with only two studies using multiple imputation. Approximately 40% of studies did not assess calibration, and approximately 60% neglected internal validation, which may have resulted in potential overfitting. Two studies relied only on external validation. None of the studies involved statistical treatment of competing risks or control subjects. The literature quality assessment is illustrated in Figure 3(High frequency predictor frequency table). Details regarding compliance with the TRIPOD-AI checklist are provided in Supplementary Appendix 4, which ranges from 2.7% to 100%. The most commonly reported items included abstracts, introductions, and sources, while blinded assessments of outcomes, missing data, risk groups, and model specifications were significantly underestimated.

## 4 Discussion

The primary goal of spinal surgery is to alleviate pain, restore function, stabilize the spinal structure, prevent neurological deterioration, and enhance the quality of life for patients. However, postoperative SSI remains a debilitating complication for patients and leads to substantial increases in medical and social costs (Hsiung et al., 2023). Effective perioperative management is essential to prevent SSI after spinal surgery, including comprehensive preoperative preparation, intraoperative antibiotic prophylaxis and irrigation with povidone iodine solution, and postoperative monitoring (Lemans et al., 2018; Tomov et al., 2015). In clinical practice, predictive models play a key role. The core is to accurately identify high-risk individuals, thereby laying a scientific basis for implementing targeted interventions and significantly improving medical efficiency and resource allocation. Given the model’s wide application, it is crucial to rigorously evaluate its effectiveness according to the principles of evidence-based medicine, which is directly related to its reliability and practical value as a clinical auxiliary tool.

This review analyzed 37 studies and identified 43 predictive models for SSI after spinal surgery. The models exhibited moderate to high predictive performance, with AUC values ranging from 0.610 to 0.991 across both internal and external validation. Specifically, four studies were externally validated, focusing on diabetes, age, operative time, albumin, BMI/obesity, smoking history, and ASA score, with AUC values ranging from 0.722 to 0.970. Overall, these studies were assessed as having a high risk of bias, with two-thirds of the studies flagged as having significant concerns about their applicability based on PROBAST-AI criteria. Adherence to the TRIPOD-AI checklist ranged from 2.7% to 100%.

SSI after spinal surgery is influenced by numerous factors (Anderson et al., 2017; Ogihara et al., 2021), including advanced age, diabetes, BMI, various somatic comorbidities, smoking, malnutrition, duration of surgery, intraoperative blood loss, and prolonged postoperative drainage time (Pennington et al., 2019). Our meta-analysis highlighted several predictors strongly associated with SSI, particularly diabetes and BMI/obesity, which aligns with the findings of Cross et al. (Cross et al., 2014). Perioperative patients are prone to abnormal blood glucose fluctuations due to pathological and psychological stress, surgical trauma, drug treatment, and nutritional support—all of which increase the risk of infection and postoperative complications. Studies (Hine et al., 2017) have indicated that the poor glycemic control in diabetic patients significantly increases the incidence of skin and soft tissue infections compared to patients with well-controlled blood glucose levels. Therefore, stabilizing perioperative blood glucose levels in diabetic patients is key to reducing complications such as postoperative infection. Our findings also support an association between higher BMI and increased SSI risk. This is consistent with most studies reporting an association between obesity/BMI and SSI (Chaichana et al., 2014; Kurtz et al., 2012). In obese patients, thicker subcutaneous fat layers necessitate deeper surgical incisions and more difficult exposure of the surgical field. The excessive stretching of the operator and the use of an electrotome can cause damage to adipose tissue. When the incision is closed, it is easy to leave dead space, and fat liquefaction and necrosis can occur after surgery, which increases the risk of wound infection (Pull ter Gunne & Cohen, 2009). Additionally, age is an important factor in the incidence of SSI after spinal surgery. Studies have indicated that the incidence of SSI and related complications, such as death and wound dehiscence, is higher in elderly patients than in the general population (Ghogawala et al., 2001). We considered that the aging of the body and the deterioration of the immune system function in elderly patients are accompanied by various underlying diseases, such as vasculopathy caused by hypertension, which reduces the local blood supply at the surgical site. In addition, heart disease reduces the heart’s ability to pump blood and reduces the blood supply to the tissues. Diminished blood flow weakens both the local immune response and tissue repair capacity, increasing the risk of infection. Smoking history is also a significant factor in developing SSI after spinal surgery. Xing et al. (Xing et al., 2013) demonstrated that after spinal surgery, smokers have a significantly higher risk of SSI than non-smokers because long-term smoking can cause vasculopathy, which leads to tissue ischemia and wound healing. This may be because long-term smokers may develop metabolic disorders related to nitric oxide, which is produced by the endothelium and acts on smooth muscle cells. It plays an important role in regulating vasomotor activity, controlling hemodynamics, and inhibiting platelet aggregation. Interestingly, smoking as a protective factor for SSI tended to be significant in multivariate analysis (Yamamoto et al., 2023). This aligns with the suspected relationship between smoking and SSI reported in previous literature (Schuster et al., 2010). It indicates that smoking may not be an independent predictor, but instead may interact complexly with other variables. Although there are related studies that indicate a correlation between hypoproteinemia and postoperative SSI, the correlation is not obvious, and may have a long half-life (Shi et al., 2021). However, Klein et al. (Klein et al., 1996) suggest that hypoproteinemia in patients with low oncotic pressure can lead to fluid loss, which can cause local wound edema and also interfere with wound healing. Concurrently, due to the increase in surgical incision exudate, bacteria around the incision and on the skin surface can easily enter the incision, thereby increasing the risk of wound infection. In addition to these preoperative factors, procedure-related factors, especially operative time and ASA classification, were identified as key predictors of SSI. A retrospective survey of 6,628 inpatients conducted by foreign scholars revealed that the risk of SSI increased with the duration of the operation (Abdul-Jabbar et al., 2012). The risk of pathogenic microorganism infection also increases with the duration of exposure to the surgical incision. The longer the operation time, the longer the tissue traction time, and the greater the degree of ischemia will be. The more inflammatory factors are released, the more easily the tissue becomes ischemic and necrotic. The ASA score was associated with an increased risk of SSI, which aligns with several previous findings (Li et al., 2019; Lieber et al., 2016). Drainage time was also an important predictor in this study, but the results were different from those of previous studies. A meta-analysis indicated that non-significant difference in the incidence of SSI was observed when wound drainage systems were used, possibly related to the inclusion of fewer data on drainage timing in this study (Tan et al., 2020).

Important insights can be drawn from the development process of the included models. In the study by Wang et al., the rigorous approach involved constructing a model using prospective data and validating it internally through 10-fold cross-validation, complemented by retrospective data for external validation. The use of external validation in the design of models effectively identified predictors of SSI after spinal surgery (Wang et al., 2021). Although the methodology was robust, their study identified postoperative albumin elevation as a predictor of SSI after spinal surgery. These conditions interact in such a way that each condition may trigger the other, underscoring the need for clear definitions and precise timing in assessments to improve the accuracy of results. Besides, the problem of insufficient sample size is frequently encountered in similar studies. The EPV metric is crucial for determining sample size adequacy, and an EPV of 20 is ideal when event rates are below 20%, with a recommended minimum of 10. For event rates between 20% and 80%, a high EPV is essential for the validity and reliability of the study (Zebley et al., 2024). Although Liu et al.’s study had a large sample size, it significantly increased the risk of bias in retrospective studies (Liu et al., 2023). Furthermore, improper handling of missing values can lead to bias and affect the reliability of the results. Most researchers choose to exclude missing data directly, but this will cause bias in the association between factors and results, affecting accuracy and resulting in wider CIs. It is suggested that selecting the appropriate treatment methods according to the specific situation of the raw data may be beneficial, such as data imputation, value deletion, maximum likelihood estimation, interpolation, and multiple imputation (Efthimiou et al., 2024). In terms of modeling techniques, Xiong et al. (Xiong et al., 2022) adopted up to seven different ML methods to determine the best model and found that the ML technique exceeded the traditional LR in accuracy. This finding is consistent with the results of four other studies (Chen et al., 2023; Liu et al., 2021; Pei et al., 2024; Wang et al., 2021). ML has advantages in managing nonlinear relationships, improving accuracy, and automating feature extraction, but it also poses challenges related to interpretability, high data requirements, and overfitting. By applying visualization techniques or local interpretation methods, the key features, weight distribution, and decision logic of the prediction model can be thoroughly analyzed, thereby enhancing the interpretability and credibility of the model in clinical scenarios, which promotes its practical application in medical decision-making (Haug & Drazen, 2023). Moreover, this study and most other studies have a key issue with the inclusion of predictors that are closely related to the outcome, such as albumin, which means that once a patient tests positive for a specific variable, the risk of death increases, and they are immediately classified as high risk.

This study highlights several key directions for future research in predictive modeling. The primary task is to optimize existing models, which can be achieved by incorporating new variables and conducting multicenter, large-sample validation. Second, the choice of predictors is crucial—we should focus on the high-frequency risk factors identified in previous studies and strictly avoid their temporal overlap with the outcome events to enhance predictive value and prevent overestimation of model performance. Additionally, methodological rigor is necessary: controlling for measurement bias in subjective variables, ensuring a sufficient time interval between predictor assessment and outcome determination, ensuring adequate sample sizes, and using appropriate methods to handle missing data. This is crucial for improving the reliability and robustness of the results. Simultaneously, it is necessary to comprehensively evaluate the model’s performance and adopt a strategy to prevent overfitting. Ultimately, this review suggests that external validation of spatial dimensions is a reliable method for assessing model validity.

## Limitations

This review has several limitations. First, the data included in the meta-analysis were only available with an OR and CI, which limits the comprehensiveness of the analysis. Second, due to methodological differences and inconsistent data reporting in the original studies, this study was unable to quantitatively integrate and analyze the overall performance of the model. Third, the literature search only covered publications in Chinese and English, which may have missed literature in other languages or informal sources (conference abstracts). Fourth, some models evaluated lacked adequate internal or external validation. This widespread lack of validation may introduce bias, affecting the reliability of research conclusions and the generalizability of models.

## Conclusion

This systematic review included 37 studies involving 43 best models. Although many studies have indicated that models constructed using different algorithms perform well regarding prediction accuracy, the best performance metrics do not necessarily imply good clinical applicability (Andaur Navarro et al., 2021). In addition, the model developed by ML for SSI can be integrated with digital platforms, enabling the creation of user-friendly web applications or platforms. These tools facilitate direct input of data to enhance understanding of disease risk. These advances support healthcare professionals by enabling early prevention and tailored, precision-based interventions.

## Data Availability

All relevant data are within the manuscript and its Supporting Information files.

## Conflicts of Interest

None

The study protocol was registered in the Prospero International Prospective Register of Systematic Reviews (CRD42024516854).

Found

None

Revision of information in the proposal

None

open access to information

Appendix 1

